# Comprehensive Evaluation and Analysis of Multi-Rod Constructs in Adult Spinal Deformity

**DOI:** 10.1101/2023.12.05.23299347

**Authors:** Moatasem M Azzam

## Abstract

Adult Spinal Deformity (ASD) is a degenerative condition of the spinal column that is categorized by sagittal imbalance, iatrogenic spinal deformity, and abnormal spinal curvature and leads to diminished quality of life. ASD is frequently repaired via neurological or orthopedic spine surgery to restore normal curvature and alignment. Surgical correction often involves the use of single, dual, or multiple-rod constructs. Multirod constructs, in particular, offer improved stability and deformity adjustment that is unmatched by single and multi-rod assemblies, but may include significant disadvantages. This review seeks to weigh the costs and benefits associated with the use of multiple rods in adult spinal deformity surgery.

## Introduction

Several studies have conveyed the effectiveness of multiple-rod constructs in ASD. These articles include case reports and literature reviews that describe multi-rod usage in surgery as effective in treating major spinal degeneration. Moniz-Garcia et al. and Guevara-Villazón et al. suggest that fusion with multiple rods results in more evenly distributed forces across the vertebral column. Bourghli et al. found little to no difference in success between single and multi-rod constructs--an indication that multi-rod constructs may not be as effective as purported. The existence of these contrasts in the literature with regards to multi-rod use led to the aim of this paper, which is to critically review the existing body of work on use of multi-rod constructs in adult deformity surgery.

## Material and Methods

A review of the literature was performed to address the following clinical question: “Are the results of multiple rods (three or more) similar to or better than those achieved with dual rods for the treatment of adult spinal deformities?”

## Literature Search Strategy

The following databases were used for screening: PubMed.gov (http://ncbi.nlm.nih.gov/pubmed), Web of Science (https://webofknowledge.com) and Scopus (www.scopus.com) to identify published articles comparing outcomes for multiple rods (MR) versus dual rods (DR) in patients adult spinal deformity (ASD). Search was not limited by dates.

For the search, the following combined search terms were used: “multiple rod OR multi-rod OR supplemental rod OR additional rod OR dual rods” AND “adult spinal deformity OR adult scoliosis OR spinal deformity OR three column osteotomies OR spine surgery”.

The search strategy was constructed after discussion and consensus of all authors. Titles found in the three databases were compared, duplicate records removed, and the remaining listings screened for inclusion by title and abstract review. Full-text manuscripts of all papers included were reviewed to ensure that all relevant papers were captured, as were all cross-referenced articles. Eligibility assessments were performed independently in a standardized manner by individual review. For further search details, please see the PRISMA flow chart in Figure 3.

## Eligibility Criteria for Study Selection

Selection criteria were as follows:

- Article published in English or Spanish.
- Article published in a peer-reviewed scientific journal.
- Article describes either a prospective or retrospective clinical trial.
- Study compares MR and DR, with MR defined as any configuration that involves more than two rods (Figure 2); and DR defined as the classic configuration of two rods connecting all the screws (see Figure 3).

**Figure 2.**
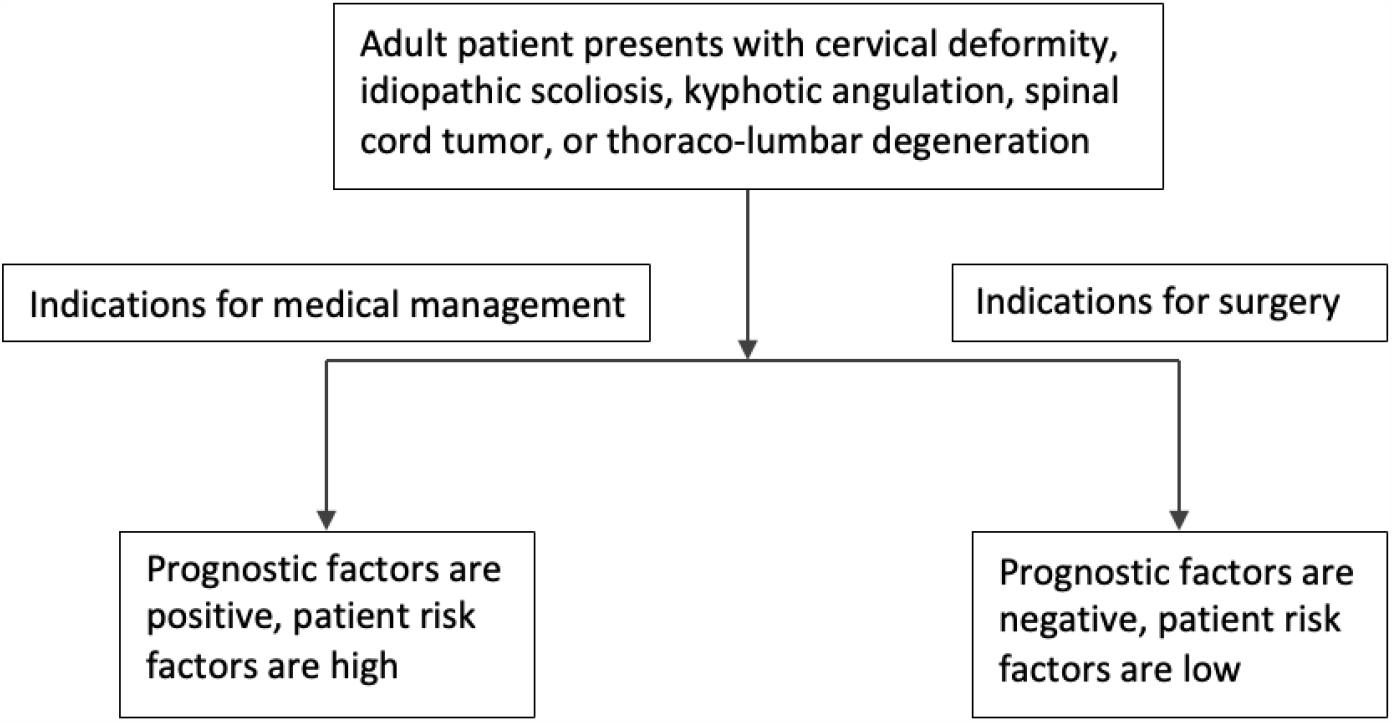
Conservative Management Versus Surgery Discussion

**Figure 3.**
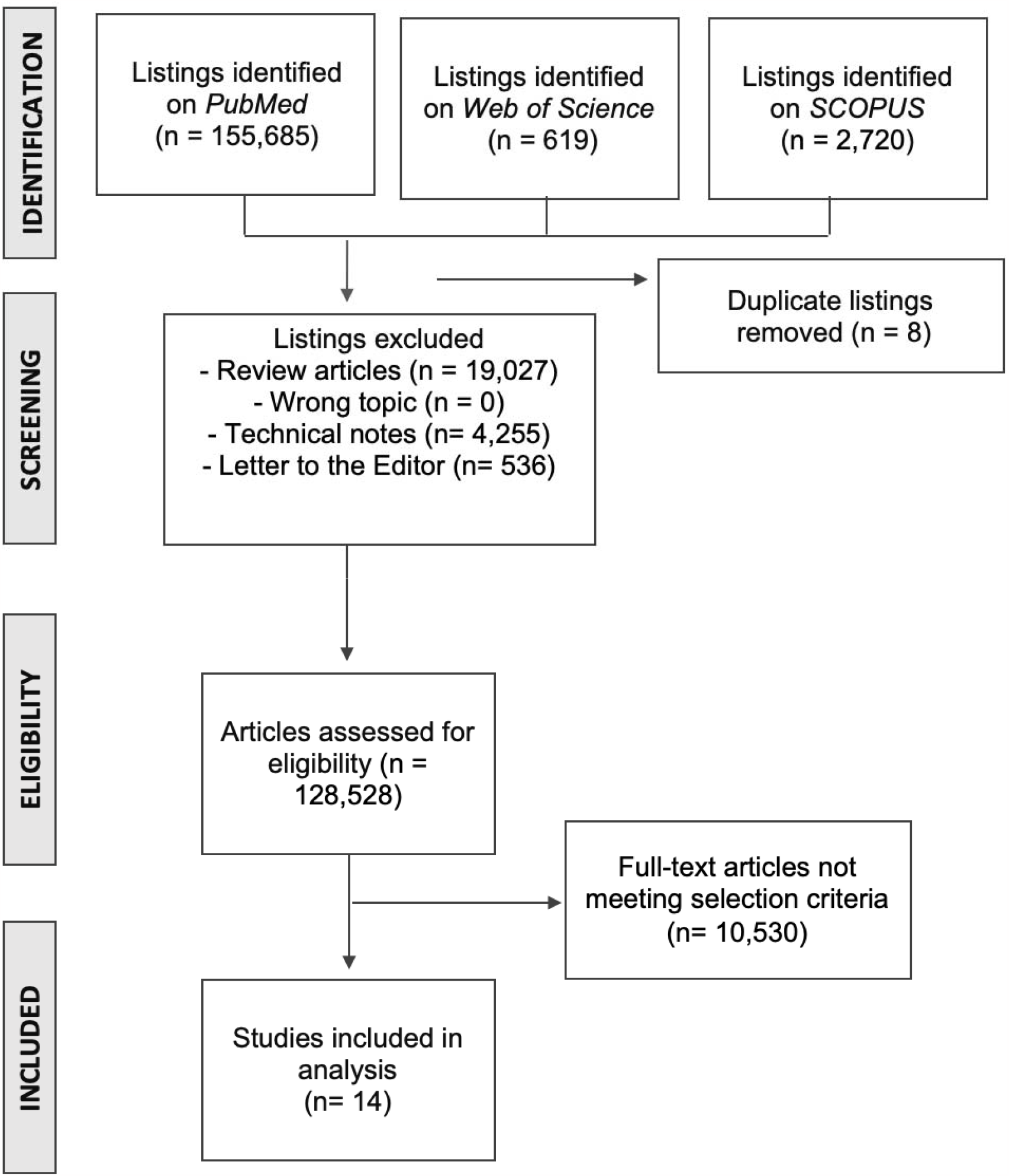
Flowchart of Identification, Screening, Eligibility, and Inclusion of articles

**Figure 5.**
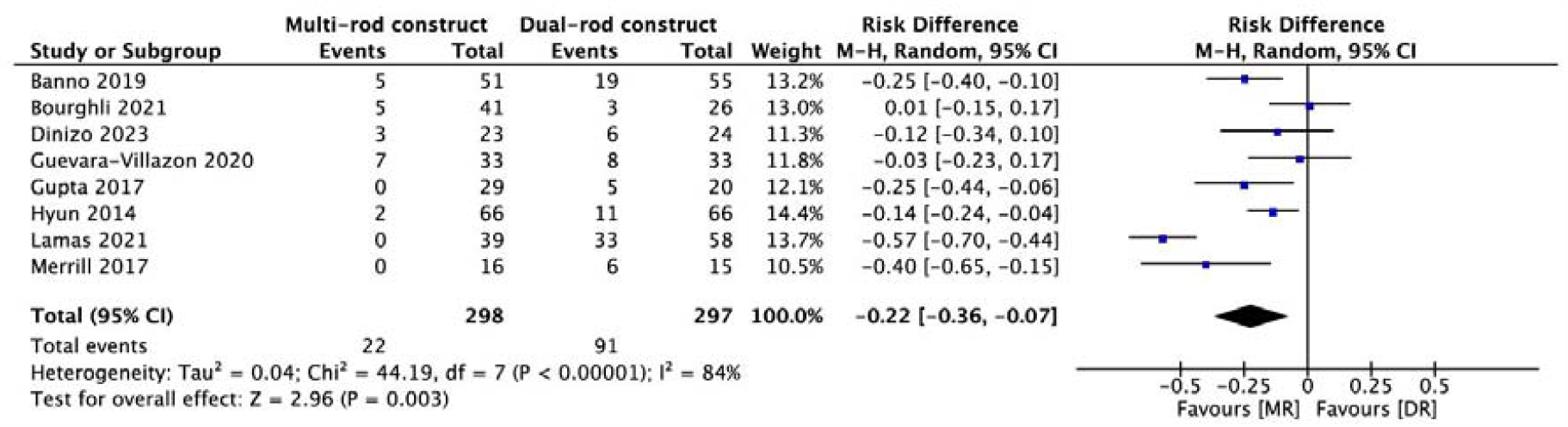
Risk Difference Between MR Versus DR
- Article reports the following data: (a) radiological, including number of rods, use of cross/connectors, rod material and its configuration (i.e. delta configuration, accessory delta); (b) surgical variables (blood loss, operative time); and (c) complications (including rod fracture, pseudoarthrosis and need for revision surgery).
- Systematic reviews and meta-analyses, letters to the Editor, book chapters, commentaries, papers with an overly high risk of bias and papers that did not meet the above inclusion criteria were excluded.

## Data extraction and analysis

After excluding all ineligible papers, the full text of each remaining article was reviewed in detail. Baseline characteristics extracted from each paper included first author, year of publication, study architecture, type of rod configuration, and the abovelisted clinical and radiological outcomes. Data were compiled and organized using Microsoft Excel 2023©.

## Methodological Quality Evaluation

All the studies were analyzed for internal validity and graded for level of evidence, in accordance with the Oxford Centre for Evidence-Based Medicine.

For risk of bias assessment ROBINS-I (Risk of bias in non-randomized studies of interventions) tool was used.

Seven sources of bias for each paper were evaluated: confounders, subject selection, classification of interventions, deviations in interventions, missing data, biased measurements and biased reporting. Finally, an overall risk of bias rating was performed as follows:

- Very Low: if all seven potential sources of bias are considered of low risk.
- Low: if no more than a single source of bias is considered of moderate risk.
- Moderate: if 2 or 3 potential sources of risk are considered moderate and none are considered of high risk.
- High: if 4 or more potential sources of risk are considered moderate or any source is considered of high risk.

**Table 1.** Bias Ratings in Accordance with ROBINS-I Tool

| **Risk of Bias in Non-Randomized Studies of Interventions** |  |  |  |  |  |  |
| --- | --- | --- | --- | --- | --- | --- |
| **Study** | **Confounding** | **Selection** | **Classification** | **Variation** | **Measures** | **Overall** |
| **Hyun et al., 2014** | Low | Moderate | Moderate | Low | High | High |
| **Banno et al., 2019** | Low | Low | Low | Low | High | High |
| **Hallager et al., 2016** | Moderate | Moderate | Low | Moderate | Low | Moderate |
| **Merrill et al., 2017** | Low | Low | Low | Moderate | High | High |
| **Lamas et al., 2021** | Low | Low | Low | Moderate | Low | Low |
| **Guevara-Villazon et al., 2020** | Moderate | Moderate | Low | Low | High | High |
| **Luca et al., 2016** | Moderate | Moderate | Low | Low | Low | Moderate |
| **Leszczynski et al., 2022** | Moderate | Moderate | Moderate | Low | Low | Moderate |
| **Jazini et al., 2021** | Low | Moderate | Low | Low | Low | Low |
| **Yamato et al., 2022** | Low | Low | Low | Moderate | Moderate | Moderate |
| **Bourghli et al., 2021** | Moderate | Low | Low | Low | Low | Low |
| **Berjano et al., 2019** | Moderate | Moderate | Low | Low | Low | Moderate |
| **Dinizo et al., 2023** | Moderate | Moderate | Low | Low | Moderate | Moderate |
| **Gupta et al., 2017** | Low | Moderate | Moderate | Low | Moderate | Moderate |

## Results

159,024 papers were identified across three databases. Two authors (MA, FH) independently evaluated the bibliographic quotes and selected the relevant abstracts. Elimination of duplicated articles and selection of eligible studies was done using Rayyan. Following search, the number of duplicates was narrowed to zero (reference system ID 542849480-9). After the application of inclusion/exclusion criteria, and removal of articles that did not meet selection criteria (Figure 3) 14 full article studies remained. Nine studies were retrospective cohort (two matched) studies that compared outcomes of dual rod construct versus multiple rod construct. Two studies involved cadaveric specimens and four studies were done via finite element analysis. The total number of operations included 346 dual rod constructs and 316 multi-rod constructs, with an addition of 11 single rod constructs and 61 lumbar Pedicle Subtraction Osteotomies (PSO).

**Table 2.**
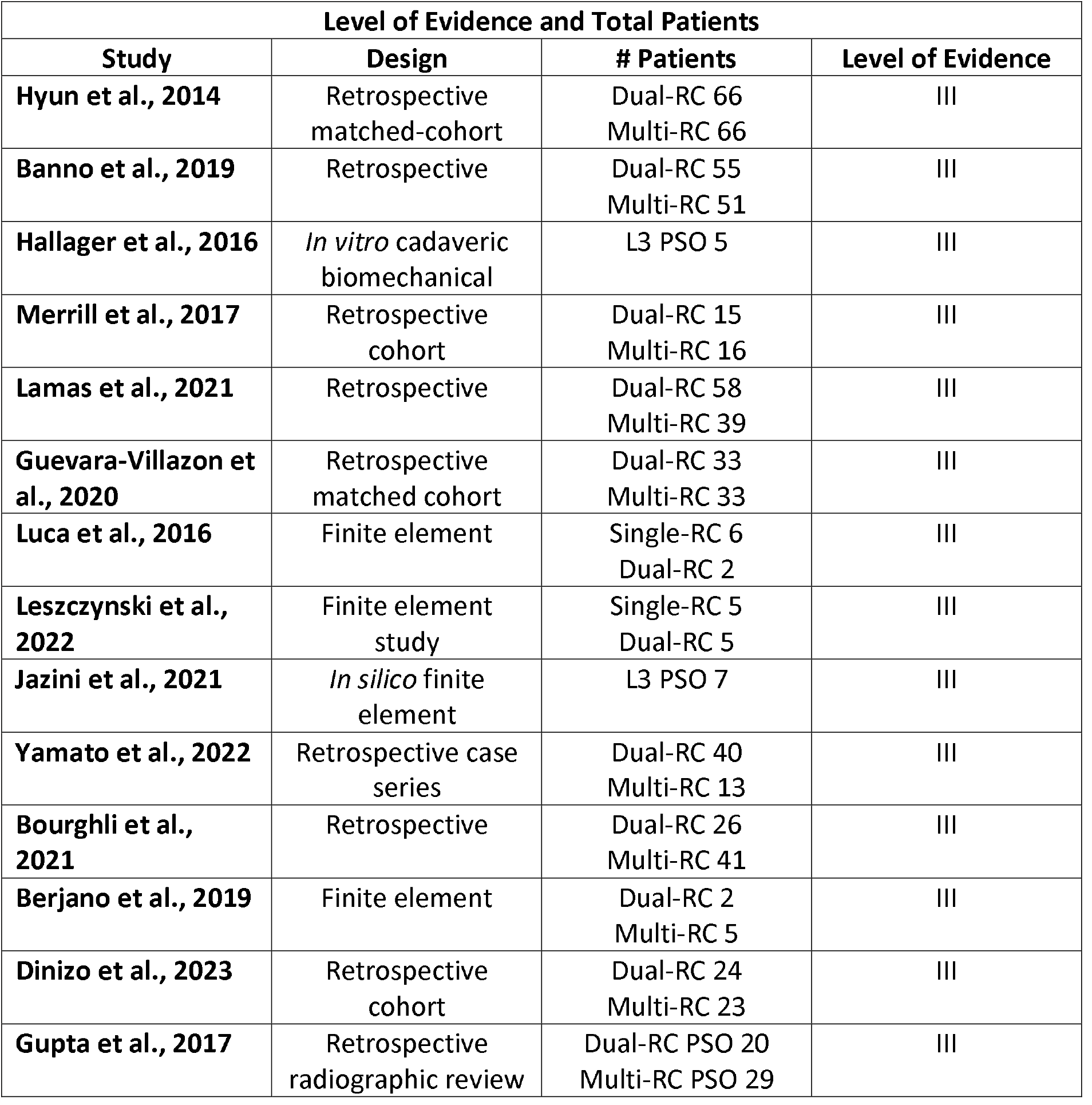
Design, Number of Patients, and Level of Evidence by Study

There were no significant differences found in range of motion following bilateral single or double rod instrumentation; however, double rod constructs were associated with higher stress reduction on the screws (Luca et al., 2016). Clinical scores improved significantly in both two- and four-rod instrumentation groups, with greater long-term improvement of lumbar lordosis and lower incidence of non-union revision surgery seen in four-rod instrumentation groups (Lamas et al., 2020). Rod strain was found to be greater with traditional inline multi-rod techniques, in comparison with Ames-Deviren-Gupta multi-rod techniques, and more asymmetric with three-rod inline technique versus more symmetric when using four-rod inline technique (Jazini et al., 2021).

Use of delta and delta-cross rods, where bends are introduced to the proximal and distal segments, lead to 45% more decreased range of motion and 48% more reduction of rod stress than satellite and accessory rods for pedicle subtraction osteotomies (PSO) (Berjano, et al., 2018). Bilateral single and double rod instrumentation showed 50 to 100% decreases in range of motion and greater rod stress found in single rod groups (Leszczynski et al., 2022). Comparison of accessory 4-rod versus primary 2-rod instrumentation and titanium versus cobalt chrome alloy rods demonstrated significant reductions of flexion-extension across all groups, with decreased relative rod strain in 4-rod groups (Hallager et al., 2016).

Rod-related complications were similar across 2-rod, multi-rod PSO, and multi-rod lumbosacral junction groups, although greater health-related quality of life scores were found in multi-rod PSO and lumbosacral junction groups (Bourghli et al., 2021). Among three groups of 55 patients who underwent two-rod operation, 16 patients who underwent three-rod instrumentation, and 35 who underwent four-rod surgery, three- and four-rod groups experienced greater incidence of complications associated with iliac and upper instrumented vertebral screws (Banno et al., 2019). Another study with two groups of 33 patients each who underwent either multi-or two-rod operations revealed no significant differences in perioperative parameters, but more frequent rod breakage and revision surgery in the two-rod group (Guevara-Villazon et al., 2019).

An evaluation of clinical results from revision surgery displayed rod fractures in 21.8% of patients at an average of 28.3 months following multi-rod construct and minimal refracture (Yamato et al., 2021). Significant differences in rod breakage between 2-rod construct and multi-rod construct groups were indicated in a study of revision for pseudoarthrosis (Hyun et al., 2014). One systematic review had insufficient evidence to indicate prevention of junctional complications after spinal fusion with multi-rod constructs of two or three-column osteotomies (Echt et al., 2021). However, comparison of a 15-patient dual-rod group and 16-patient multi-rod group exhibited zero incidence of rod fracture in the multi-rod group versus 40% incidence in the dual-rod group, predominantly in the lumbosacral junction (Merrill et al., 2017).

Highlighted first is the importance of this paper as guidance in order to define when the use of multi-rods is justified and when not. Multiple-rod constructs are used in the surgical repair of adult spinal deformities, often to mitigate the risk of postoperative revision due to rod fracture. An operation for abnormal spinal curvature requires the use of rods to stabilize vertebral segments. In the case of spinal fusion surgery, rods may also be used to connect screws and therefore limit movement across discs. The use of multiple rods in spine surgery provides additional stability to the vertebral column and decreases the tension absorbed by each individual rod (Hallager et al., 2016). However, there are both advantages and disadvantages to the use of multi-rod constructs as opposed to traditional single or dual-rod approach.

Studies that have examined the use of multi-rod constructs in spinal surgery conclude that multi-rod construct installment is associated with advanced correction of spinal malcurvature over single-rod construct insertion. The incidence of intergroup multi-, two-, and single-rod fractures differs across studies, where some find no significant differences in complications associated with the number of rods (Bourghli et al., 2021) and others pointed towards greater prevalence of degenerative fragmentation for two-rod installations (Guevara-Villazon et al., 2019). This evidence suggests no outright advantage through the use of either strategy when scrutinized for rate of complications, and instead that the ideal outcome with minimal revision may be achieved through careful contemplation of how individual patient spine can be augmented with chosen apparatus.

Rod fractures can be a frequent occurrence following their use in spine surgery, arising in anywhere from 21.8% (multi-rod) to 40% of patients who undergo fixation (Yamato et al., 2021; Merrill et al., 2017). In addition to the particular surgery and number of rods, incidence of deterioration may vary according to the alloy, dimensions, or shape used. Examples include stainless steel, titanium, or cobalt-chromium; 5mm, 5.5mm, or 6 mm; and delta, satellite, or accessory rods (Lamas, et al., 2020; Berjano, et al., 2018; Luca et al., 2016). Proper selection of instrumentation is critical, as this can affect previously addressed factors as well as operation costs. Surgeons who propose increasing the number of rods as a strategy for reducing the rates of rod fractures should use these to assess the risk of breakage.

These results suggest that multiple rod constructs provide greater relief from symptoms, as compared to single-rod constructs. The use of more rods is associated with greater improvement of spinal deformity and lower incidence of revision surgery as a consequence of rod strain and fracture (Banno et al., 2019; Bourghli et al., 2021; Guevara-Villazón et al., 2020; Hyun et al., 2014; Lamas et al., 2021). In addition, the degree of benefit can be seen as a function of traditional versus non-traditional strategies or techniques, symmetric versus asymmetric placement, and the type of material used. This study assessed each of these parameters to determine the effectiveness of multi-rod use and found differences according to each. Moreover, this study delineated the additional benefits and some disadvantages brought about by increased number of rods inserted that mostly concern decreased range of motion.

The statistical analysis revealed favorable correction associated with multi-rod surgery, as opposed to dual-rod surgery, with five out of eight studies showing a significant difference in outcomes. Three out of eight studies showed no significant difference between multirod surgery and dual-rod surgery. The 95% confidence intervals of statistical significance were [-0.40, -0.10] (Banno 2019), [-0.44, -0.06] (Gupta 2017), [-0.24, -0.04] (Hyun 2014), [-0.70, -0.44] (Lamas 2021), [-0.65, -0.15] (Merrill 2017). Total 95% confidence interval of the risk difference between MR and DR was [-0.36, -0.07], from the eight studies out of fifteen studies that were included. Only overall measures were compared due to the large heterogeneity of between papers (p < 0.00001).

Indications for multi-rod surgery of ASD include degenerative changes, scoliosis of known or unknown etiology, congenital or post-traumatic deformity, and post-intervention spinal deterioration. The decision to perform a multiple rod surgery specifically, in lieu of single or dual-rod surgery, varies by physician and depends on several variables such as patient risk factors, degree of spinal angle regression, and the number of vertebrae in-need of fusion (a variable associated with the magnitude of deformity). Adult idiopathic scoliosis, proximal or distal junctional kyphosis, spinal cord tumor, and cervical deformity are typical conditions that can be treated with multiple rod fusion of vertebral segments.

The evidence suggests that if there does not exist a severe deformity, high degree of instability, or large number of vertebral levels that require fusion, the surgeon should not proceed with the operation and default to conservative medical therapy. Further evaluation should uncover if there is a high risk of rod fracture and increased likelihood of implant failure, and if not, to proceed with single rod construct. Experience with the presenting adult spinal deformity and use of multiple-rod constructs should be the surgeon’s final consideration of whether the patient is best served by undergoing an operation that requires a dual-rod or multiple-rod construct.

Multi-rod assemblies in spine have been shown to provide certain benefits that include decreased incidence of implant fracture, increased deformity reduction, and improved stability of vertebrae with construct. In concert with higher complexity, the risks associated with multi-rod implantation may also involve decreased range of motion, increased operative time, and increased blood loss. Although no significant difference was found in range of motion between patients treated with bilateral single or double rod instrumentation, there was an effect mediated by the shape of rod. This finding presents an important consideration for rod selection in conjunction with the actual number of rods. The comparison across non-single rod groups illustrated that range of motion, measured via spinal flexion and extension, decreased across both groups, therefore the surgeon should appraise the number of rods in tandem with rod material that should be used when evaluating individual patient characteristics.

Studies that have examined the use of multi-rod constructs in spinal surgery conclude that multi-rod construct installment is associated with advanced correction of spinal malcurvature over single-rod construct insertion. The incidence of intergroup multi-, two-, and single-rod fractures differs across studies, where some find no significant differences in complications associated with the number of rods (Bourghli et al., 2021) and others pointed towards greater prevalence of degenerative fragmentation for two-rod installations (Guevara-Villazon et al., 2019). This evidence suggests no outright advantage through the use of either strategy when scrutinized for rate of complications, and instead that the ideal outcome with minimal revision may be achieved through careful contemplation of how individual patient spine can be augmented with chosen apparatus.

Spine surgery has seen a rise in the use of multi-rod constructs due to the positive outcomes associated with their utility in complex cases. The advantages of multi-rod models encompass prevention of hardware failure and consequently enhanced fusion, facilitation to correct more considerable or pronounced spinal deformities, and reduced need for revision subsequent to the aforementioned improvements. However, neurological and orthopedic spine surgeons should not lose sight of possible downfalls.

The disadvantages of multi-rod frameworks include added complexity associated with a larger number of rods and related complications, increased intra- and post-operative blood loss, and longer time of surgery that may compound the rates of post-surgical shock, infection, or thrombosis. However, multiple rod constructs used in spinal surgery can improve the rates of column fracture, the degree of deformity correction, and the strength of vertebral instrumentation. Such results and evidence thereof may serve as landmarks for future research on implementation of multiple rods in spine adjustment.

Notable limitations of this systematic review are associated with the lack of prospective studies on comparison of dual- and multiple-rod constructs, the low number of studies that were included after meeting eligibility criteria, and variability in the quality of studies included. The deficit of prospective investigations on multi-rod use in adult spinal deformities may be attributed to the relatively greater length of time needed to follow-up with recipients of invasive spine surgery. In addition, the seeming absence of articles covering the topic of interest can be ascribed as a consequence of the standards for selection into review. These standards excluded all studies that were not prospective or retrospective by design, and that did not directly compare dual- and multi-rod constructs. Studies ranged from low to high risk of bias upon ROBINS analysis, but all remained included due to the high level of evidence present in each.

## Conclusion

This study found that the use of multi-rods for adult spinal deformity appears to be protective against rod fracture and pseudoarthrosis. Numerous studies have scrutinized the advantages and disadvantages of multiple rod assemblies in scoliosis, kyphosis, and lordosis treatment. The primary advantages of a multiple rod approach include decreased likelihood of post-surgical complications and longer-term adjustment with regards to the angulation of concern. Disadvantages can include decreased range of motion, increased operative time, and increased blood loss (Merrill et al., 2017). These considerations must be weighed pre-operatively in order to decide whether a multiple or traditional construct is more suitable. Otherwise, the surgeon risks greater intraoperative challenges and worse clinical outcomes.

## Data Availability

All data produced in the present study are available upon reasonable request to the authors

